# Expression of Exosome Biogenesis Genes is Pervasively Altered by Aging in the Mouse and in the Human Brain During Alzheimer’s Disease

**DOI:** 10.1101/2021.08.02.21260877

**Authors:** Daniel S. Lark, Thomas J. LaRocca

## Abstract

Extracellular vesicles (EVs) like exosomes are secreted by numerous cell types in a variety of tissues. EVs have been implicated in both aging and age-related disorders like Alzheimer’s disease (AD). However, how aging and AD affect EV biogenesis within and across cell types is poorly understood. Moreover, cells acquire characteristics based on tissue niche, but the impact of tissue residence on cell type EV biogenesis is unknown.We explored the *Tabula Muris Senis*, Mayo RNA-seq and ROSMAP data sets to characterize the cell and tissue-specific effects of aging and AD on genes involved in EV biogenesis. Specifically, we examined the age-dependent expression (age coefficient) of genes involved in EV biogenesis (22 genes), EV cargo (3 genes) and senescence (5 genes). Of the 131 cell populations (cell type x tissue) studied, 95 have at least one EV biogenesis gene impacted by age. The most common gene increased by age was charged multivesicular body protein 2A (CHMP2A) (54 cell populations). The most common gene decreased by age was syndecan binding protein (SDCBP) (58 cell populations). The senescence-associated genes cyclin-dependent kinase 1A (CDKN1A) and CDKN2A were not related to changes in CHMP2A and SDCBP and were altered by age in fewer cell populations. Finally, individuals with AD had decreased CHMP2A and increased SDCBP expression, opposite of what is observed with aging in the absence of diagnosed neurological disease. These findings indicate that age modifies exosome biogenesis gene expression in many cell populations mostly independent of senescence, and may be further altered in AD.

## Introduction

Advancing age is a primary predictor for the development of chronic disease [1]. The mechanisms linking aging to chronic disease are incompletely defined, but recent work indicates an important role for extracellular vesicles (EVs) like exosomes. Exosomes are lipid-encapsulated particles ranging from 50-200 nm in diameter [2]. Exosome biogenesis occurs through an endosomal process coordinated by a series of endosomal sorting complexes required for transport (ESCRTs) [3]. To date, very little is known regarding how the ESCRT pathway changes during aging. In fact, to our knowledge, the only evidence linking the ESCRT pathway to aging comes from recent work demonstrating that the ESCRT-II [4] and ESCRT-III [5] complexes are important for replicative lifespan in yeast.

Recent work showing that exosomes from young donors [6] or young mesenchymal stem cells [7] extend lifespan in rodents demonstrates the potential for using EVs as an anti-aging therapy. However, somewhat in contrast, there also is growing evidence that senescence leads to an increase in total exosome secretion [8-10], although the mechanism(s) involved and the function of these EVs have not yet been described. While senescence is a hallmark of aging in some cell types, senescence and aging are not mutually inclusive [11]. It is not known if, and to what extent, aging itself affects the biogenesis and secretion of exosomes (i.e., in the absence of senescence). Additionally, it is not currently known whether EV biogenesis is related to classic age-related disorders like Alzheimer’s disease (AD). Elucidating the impact of aging on exosome biogenesis will not only improve our understanding of the basic biology of aging, but may also improve the design and implementation of exosome-based therapeutics by identifying the cell types, and molecular mechanisms, most affected by aging.

Recent advances in single cell RNA sequencing (scRNA-Seq) have provided new opportunities to study the heterogeneity of aging within and between tissues and cell types that would otherwise be impossible or impractical. For example, the *Tabula Muris Senis* is a scRNA-Seq dataset comprising 131 different cell populations (type x tissue) from 22 different tissues across the lifespan of the C57BL/6JN mouse [12]. Investigators from the Tabula Muris consortium generated age coefficients to describe the relationship between age and gene expression across the lifespan. Here, we analyzed age coefficients from the *Tabula Muris Senis* to define how aging impacts exosome biogenesis pathways/gene expression, and the association of these changes with senescence and exosome cargo. We then compared our findings to publicly available RNA-Seq data on human AD. Our analysis: 1) illustrates that aging pervasively alters exosome biogenesis gene expression in the mouse, 2) identifies two novel aging-associated genes/transcripts involved in exosome biogenesis via the ESCRT pathway and 3) implicates exosome biogenesis as an impacted process in the brain of human AD patients.

## Materials and Methods

The methods used to obtain and process data for the *Tabula Muris Senis* are described here [12]. In brief, tissues were harvested from mice between 1 and 30 months of age. Single cells were isolated and scRNA-Seq was performed. Age coefficients were generated in the *Tabula Muris Senis* using a linear model [13] where age was a numerical value and coefficients were adjusted for age and technology. We obtained differential gene expression data described in the *Tabula Muris Senis* from the Gene Expression Omnibus website (Accession number: GSE149590). Data on human AD were obtained from the Mayo RNA-seq study and the Rush Religious Order Study/Memory and Aging Project (ROSMAP) scRNA-Seq study, both available on the Synapse database (Accession numbers: syn3163039 and syn18485175).

The primary gene set of interest was the Gene Ontology category “extracellular exosome biogenesis” GO: 0097734. In addition, CD9, CD63 and CD81 were chosen because these genes encode established exosome cargo proteins [2]. Finally, Cyclin-dependent kinase 1A (CDKN1A),CDKN2A, Interleukin-6 (IL-6), Interleukin-1β (IL-1β), CCL2 and tumor necrosis factor (TNF) were chosen based on their established roles in senescence [14]. Age coefficient data was filtered based on an adjusted p-value of < 0.01. All figures were generated using GraphPad Prism software.

## Results and Discussion

To identify exosome biogenesis genes/transcripts altered by aging, we calculated the number of cell populations (population = cell type x tissue; n=131) across all tissues with significantly altered expression with age (**Figure 1A)**. The ten most commonly decreased (**Figure 1B; top**) or increased (**Figure 1B; bottom**) genes/transcripts with age are shown. Age coefficients for every cell population are provided in **Supplemental Materials**. From this analysis, we found that 95 different cell populations have at least one exosome biogenesis gene altered by aging. In general, genes more frequently decreased in expression with age. Syndecan binding protein (SDCBP; also known as Syntenin-1) was the exosome biogenesis gene most commonly decreased with age. Charged multivesicular body protein 2A (CHMP2A) was the exosome biogenesis gene most commonly increased with age. Importantly, recent reports suggest that SDCBP may be a “universal biomarker” of exosomes [15], and CHMP2A has been shown to decrease with senescence [16] (although its exact function in this context is unknown). These results demonstrate that aging pervasively impacts the expression of exosome biogenesis genes across many cell types and tissues.

**Figure 1.**
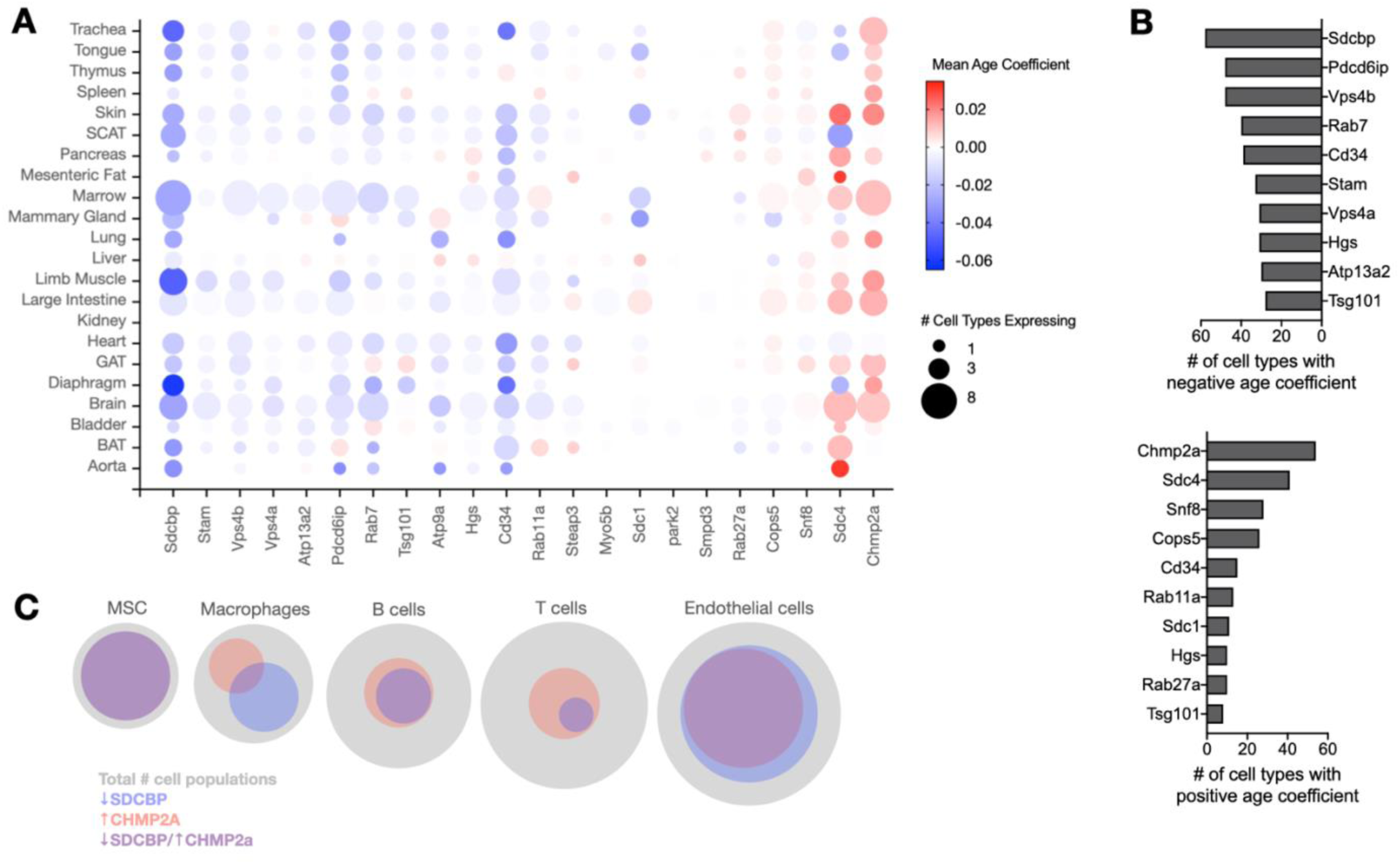
Pervasive age-related changes in exosome biogenesis gene expression in numerous cell/tissue types. **(A)** Dot plot showing number of cell populations and age-related expression changes (age coefficients) for exosome biogenesis genes across all tissues in the *Tabula Muris Senis* dataset. N=131 cell populations. **(B)** Exosome biogenesis genes ranked based on the number of cell populations displaying decreased (top) or increased (bottom) expression with age. **(C)** Proportion of cell populations with decreased SDCBP and/or increased CHMP2A (key exosome biogenesis genes) expression among the most common cell types in the *Tabula Muris Senis*.

We next asked whether decreased expression of SDCBP and increased expression of CHMP2A with aging are commonly observed in the five most abundant cell types in the *Tabula Muris Senis* (endothelial cells, T-cells, B cells, macrophages and mesenchymal stem cells). As shown in **Figure 1C**, the majority (5/6) of mesenchymal stem cell populations displayed a concurrent increase in CHMP2A and decrease in SDCBP with age. Multiple (6/14) endothelial cell populations also displayed both an increase in CHMP2A and a decrease in SDCBP. By comparison, macrophages, B cells and T cells showed some, but infrequent, co-occurrence of decreased SDCBP and increased CHMP2A expression. These findings indicate that aging preferentially affects gene expression of CHMP2A and SDCBP in mesenchymal stem cells and endothelial cells, and less commonly in immune cell populations.

Senescence is a hallmark of aging and is known to promote the release of small EVs like exosomes [8-10]. Others have shown that SDCBP gene expression is increased in senescent cells compared to proliferating cells in culture [17], and that senescence decreases CHMP2A expression [16]. However, our analysis generally suggests an age-related decrease in SDCBP expression and increase in CHMP2A expression. Thus, we sought to better understand the cell population-specific relationship between changes in expression of CHMP2A and SDCBP and senescence-associated genes. Expression of the senescence-associated genes CDKN1A, CDKN2A, IL-1β, IL-6, CCL2 and TNF was only increased in 36, 9, 17, 20, 35 and 6 cell populations (out of 131), respectively (**Figure 2A; right**). Fewer cell populations displayed a decrease in these same genes (**Figure 2A; left**). Therefore, at a glance, the exosome biogenesis genes SDCBP, CHMP2A, syndecan-4 (SDC4), programmed cell death 6 interacting protein (PDCD6IP aka ALIX), vacuolar sorting 4 homolog B (VPS4B), Rab7 and CD34 were all more commonly altered by age than any of these senescence markers.

**Figure 2.**
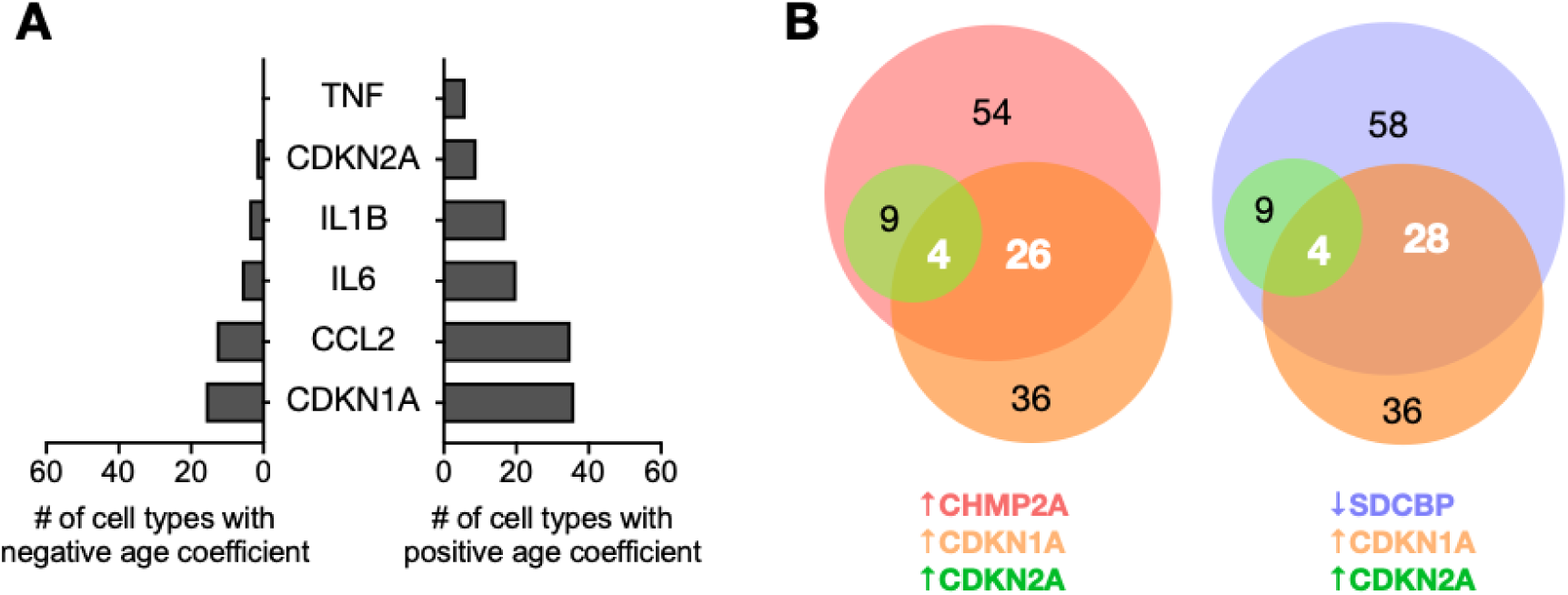
EV biogenesis genes are more responsive to aging than senescence-associated genes. **(A)** Number of cell populations from the *Tabula Muris Senis* dataset with decreased (left) and increased (right) expression of senescence genes with age. N=131 cell populations. **(B)** Proportion of cell populations with concurrent age-related changes in the expression of CHMP2A (left) or SDCBP (right) and the senescence genes CDKN1A and CDKN2A. Black numerals represent the total number of cell populations with the specific gene expression pattern with aging. White numerals represent the number of cell populations that overlap with other gene expression patterns.

Co-occurrence analysis (**Figure 2B**) indicated that only 48% of cell populations with decreased SDCBP expression (28 out of 58) or increased CHMP2A expression (26 out of 54) also displayed an increase in CDKN1A. Furthermore, only 16% of cell populations with decreased SDCBP expression (9 out of 58) and 17% of cell populations with increased CHMP2A expression (9 out of 54) also displayed an increase in CDKN2A. Collectively, our analysis strongly suggests that aging alters exosome biogenesis genes in a wide variety of cell populations and can occur independent of classic markers of senescence (i.e., CDKN1A and CDKN2A). Further, this finding suggests that distinct cellular alterations in exosome biogenesis occur during healthy aging (prior to the onset of pathological/senescence-associated aging).

Could the age-related changes in EV biogenesis gene expression we observe in mice be relevant to human aging/disease? To investigate this possibility, we examined EV biogenesis gene expression in brain aging and AD, a quintessential disease of aging [18]. Exosomes have been implicated in the etiology of AD via trafficking of tau [19, 20], and silencing of CHMP2A causes the propagation of tau seeds [21], which lead to tau tangles that are associated with AD pathology [22]—but a link to aging has not been established. Because tau accumulates even during normal aging [23], we first examined whether the aging brain is characterized by altered EV biogenesis gene expression in the *Tabula Muris Senis*. We found that seven of the nine cell types in the brain displayed age-related changes in CHMP2A expression (**Figure 3A**). CHMP2A expression was increased in six of these cell types, consistent with the general response of CHMP2A to aging, but CHMP2A expression was decreased with age in neurons. In contrast, consistent with its general response to aging, SDCBP expression was decreased in five of nine brain cell types but not increased in any brain cell type. To determine if these changes in EV-associated genes/transcripts are relevant to AD, we next examined temporal cortex tissue gene expression data from the Mayo RNA-Seq study and scRNA-seq data from the ROSMAP study. In post-mortem cortex samples from the Mayo RNA-Seq study, individuals with diagnosed AD displayed numerous alterations in exosome biogenesis genes/transcripts (**Figure 3B**) compared to neurotypical individuals. Many of these gene expression patterns were opposite of the age-related changes observed in the *Tabula Muris Senis* (e.g., decreased CHMP2A and increased SDCBP expression). Interestingly, scRNA-seq data from the ROSMAP study confirmed these AD-related changes and suggested they may be: 1) especially pronounced in neurons and microglia; and 2) progressively increased with pathology (**Figure 3C**). These data collectively demonstrate that AD is associated with gene expression changes in CHMP2A and SDCBP that oppose those observed with normal aging, and they could be consistent with a model in which age-related changes in CHMP2A/SDCBP expression in neurons contribute to the accumulation of tau and the glial cell activation that are thought to contribute to AD pathology.

**Figure 3.**
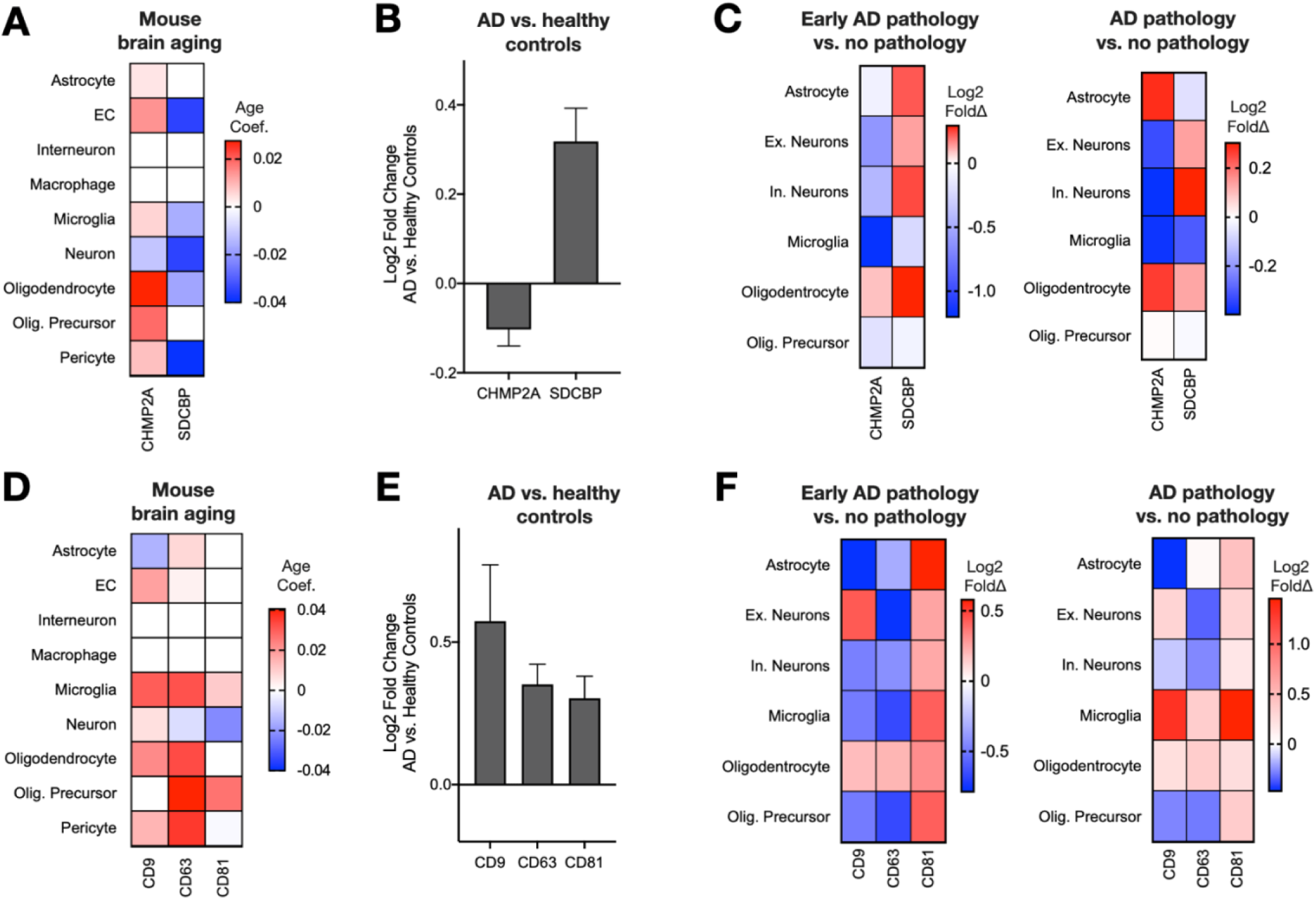
Brain aging and Alzheimer’s disease-associated changes in exosome biogenesis and cargo gene expression. **(A)** Age-related changes (age coefficients) in expression of the key exosome biogenesis genes SDCBP and CHMP2A in brain cell types in the *Tabula Muris Senis* dataset. **(B)** Differential expression of SDCBP and CHMP2A in bulk brain samples of Alzheimer’s disease patients compared to neurotypical controls from Mayo RNA-seq dataset. **(C)** Differential expression of SDCBP and CHMP2A in different brain cell types with increasing Alzheimer’s disease pathology from the ROSMAP scRNA-seq dataset. **(D)** Age-related changes in expression of key exosome cargo genes in brain cells in the *Tabula Muris Senis* dataset. **(E)** Differential expression of exosome cargo genes in bulk brain samples from Alzheimer’s disease patients compared to neurotypical controls in the Mayo RNA-seq dataset. **(F)** Differential expression of exosome cargo genes in different brain cell types with increasing Alzheimer’s disease pathology from the ROSMAP scRNA-seq dataset.

Recent work has demonstrated that exosome cargo is also altered in patients with AD [24]. In the *Tabula Muris Senis*, we found that microglia have increased expression of CD9, CD63 and CD81 with age (**Figure 3D**), whereas neurons have increased expression of CD9 but decreased expression of CD63 and CD81 with age. In the Mayo RNA-Seq study (bulk brain RNA-seq), patients with AD displayed increased expression of CD9, CD63 and CD81 compared to neurotypical controls (**Figure 3E**), and ROSMAP scRNA-seq data indicated that this could be explained by pathology-associated expression increases in microglia (**Figure 3F**), which are highly abundant in the brain. This differential expression of CD63 in microglia (increased) and neurons (decreased) is interesting since CD63 identifies a unique population of neuroprotective microglia in the 5xFAD mouse model of AD [25]. Additionally, circulating blood amyloid-beta bound to CD63^+^ exosomes has been recently shown to better predict AD than unbound circulating amyloid-beta [26]. These results indicate that gene expression changes in exosome biogenesis in the aging brain may contribute to the development and progression of AD.

Overall, our findings demonstrate that advancing age causes pervasive alterations in exosome biogenesis genes. The two most commonly altered exosome biogenesis genes are CHMP2A and SDCBP, which have not been previously implicated in aging. We also show that age-related changes in exosome biogenesis can occur independent of classical markers of senescence, and that AD dramatically impacts exosome biogenesis gene expression in the brain, including CHMP2A, which has been recently implicated in the regulation of tau trafficking. These findings offer new insight into the relationship between aging and exosome biogenesis within and across cell types and the potential relevance of these events to neurodegenerative diseases like AD.

## Data Availability

All data used in this paper are already publicly available. Links to data sets are provided in the manuscript.

## Notes

### Competing Interest Statement

The authors have declared no competing interest.

### Funding Statement

No external funding was used for this study.

### Author Declarations

Data on human AD were obtained from the AD Knowledge Portal (https://adknowledgeportal.org) and originally generated by: 1) the Mayo Clinic Alzheimer's Disease Genetic Studies using samples from the Mayo Clinic Study of Aging, the Mayo Clinic Alzheimer's Disease Research Center, and the Mayo Clinic Brain Bank (Mayo RNA-seq study, ethical approval by the Mayo Clinic Research Executive Committee); and 2) the Rush Alzheimer's Disease Center, Rush University Medical Center (ROSMAP study, ethical approval by the Rush University Medical Center). All data were accessed on the Synapse server under an approved data use certificate (TJL).

